# Clozapine metabolism is associated with Absolute Neutrophil Count in individuals with treatment-resistant schizophrenia

**DOI:** 10.1101/2021.02.01.21250894

**Authors:** Isabella Willcocks, Sophie E. Legge, Mariana Nalmpanti, Lucy Mazzeo, Adrian King, John Jansen, Marinka Helthuis, Michael J. Owen, Michael C. O’Donovan, James T.R. Walters, Antonio F. Pardiñas

## Abstract

**AIM:** To investigate the relationship between clozapine concentration and neutrophils in a European cohort of long-term clozapine users.

**METHODS:** Pearson’s Correlation and Linear Regression analyses were applied to a subset of the CLOZUK2 dataset (N = 208) to assess the association between Absolute Neutrophil Count (ANC) and plasma clozapine concentration. Norclozapine and the metabolic ratio between clozapine and norclozapine were also investigated, along with SNPs associated with clozapine metabolism

**RESULTS:** Association between ANC and plasma clozapine concentration was found to be significant in a linear regression model (β = −1.41, p = 0.009), with a decrease in ANC of approximately 141 cells/mm3 for every 0.1 mg/litre increase in clozapine concentration. This association was attenuated by the addition of the metabolic ratio, which was significantly negatively correlated with ANC (β=-0.69, p=0.021). In a further regression model, three SNPs previously associated with norclozapine plasma concentrations and clozapine/norclozapine ratio were also found to be significantly associated with ANC: rs61750900 (β=-0.410, p=0.048), rs2011425 (β=0.450, p=0.026) and rs1126545 (β=0.330, p=0.039)

**CONCLUSION:** ANC was found to be significantly negatively associated with plasma clozapine concentration. Further investigation has suggested that the relationship is mediated by the clozapine/norclozapine ratio, and potentially moderated by genetic variants with effects on clozapine metabolism

## Introduction

Schizophrenia is typically a severe, chronic neuropsychiatric disorder affecting an estimated 0.7% of the global population (McGrath et al., 2008). Although antipsychotic medications are generally effective, 20-30% of patients are classified as having ‘Treatment-Resistant’ Schizophrenia (TRS), defined as a lack of symptomatic relief following trials of at least two antipsychotics at an adequate dose for a reasonable time (Howes et al., 2017). For these individuals, treatment options are often limited to a single medication; clozapine, the first atypical antipsychotic drug developed, and the only drug currently available with proven efficacy in TRS. Clozapine has also been shown to have robust effects on preventing suicide (Meltzer, 2011) in this patient subgroup. However, although an estimated two-thirds of patients with treatment-resistant symptoms would respond beneficially to clozapine (Meltzer, 2011), it is still widely under-prescribed, in part due to the risk of serious adverse effects (De Fazio et al., 2015). One of these, affecting 0.38-0.8% of patients, is agranulocytosis, a potentially fatal condition in which there is a significant decrease in granulocytes, the most abundant form of which are neutrophils (Alvir et al., 1993). To minimise the risk of agranulocytosis, regular haematological monitoring is required for safe and effective treatment with clozapine. Clozapine-induced agranulocytosis is most likely to develop within the first 18 weeks of treatment (Atkin et al., 1996), during which time patients’ white blood cell levels are measured weekly and treatment stopped if agranulocytosis is detected or monitoring shows a sustained but milder decrease of white blood cells (neutropenia). This need for intensive physical health monitoring is an additional burden for the use of clozapine, and has been indicated by service users as a reason for discontinuing treatment (Legge et al., 2016).

The exact relationship between clozapine dose and ANC remains poorly understood, although several mechanisms have been proposed to argue that clozapine might even have a dose-dependent effect on ANC (Vaquero-Baez et al., 2019), which could, in turn, lead to neutropenia. One such mechanism is the production of a reactive nitrenium ion following the bioactivation of either clozapine or one of its primary metabolites (Liu and Uetrecht, 1995). In turn, this short-lived but highly reactive intermediate may act as a hapten leading to the destruction of neutrophils via an immune response, or it may disrupt neutrophil function by binding to crucial cellular proteins (Pirmohamed and Park, 1997). This effect could potentially be enhanced if an infection occurs, since inflammatory processes are known to increase clozapine plasma concentrations (de Leon et al., 2020; Siskind et al., 2020). Complicating the matter further is the fact that there is substantial variation of plasma clozapine concentration between individuals who have been prescribed the same dose; with modifying factors including age, weight, smoking and likely genomic variation affecting enzymes involved in clozapine metabolism (Olsson et al., 2015). It is important to better understand the link between clozapine doses, clozapine metabolism and neutrophil counts for several reasons. First, because it would allow for the evaluation of the effectiveness of interventions aimed at lessening the potential detrimental impacts of clozapine on the immune system, such as those proposed in response to the COVID-19 pandemic (Pandarakalam, 2020; Siskind et al., 2020). Additionally, in line with recent research on underrepresented populations (Legge et al., 2019), improved comprehension of the biology behind potential adverse effects can help refining current dosing and titration protocols, therefore increasing the number of TRS patients who can be safely prescribed the medication and benefit from its therapeutic effects.

Recently, a study found a negative association between plasma clozapine concentration and neutrophil count (Vaquero-Baez et al., 2019), reporting an R^2^=0.447 from multiple regression analyses, conventionally considered a “large” effect (Cohen, 1992). However, the study was arguably small, including only 41 patients. Motivated to test these findings on a larger cohort, we used similar statistical methods on the UK-based CLOZUK2 study (Pardiñas et al., 2018) of people with TRS. We also extended the design to incorporate genetic data, specifically genetic variants that have been recently found to be associated with clozapine metabolism (Pardiñas et al., 2019).

## Methods

### Samples, Neutrophil Count and Clozapine Concentration Data

ANC, clozapine concentration and genetic data were obtained as part of the CLOZUK2 study. Full details of recruitment and quality control of the samples are provided in Pardiñas et al. (2018). Curation of ANC data and assessment of genetic ancestry are described in Legge et al. (2019), while curation of the longitudinal clozapine concentration data is described in Pardiñas et al. (2019).

### Inclusion Criteria

The dataset was restricted to individuals of European ancestry who had a plasma clozapine measurement (mg/L) within 21 days of an ANC (cells/mm^3^) measurement. For any individual who had more than one clozapine plasma concentration sample within that window, the measurement closest to the ANC was selected. The clozapine/norclozapine ratio (“metabolic ratio”) was calculated for each individual, and anyone with a ratio greater than 3 and less than 0.5 was excluded (n = 19), in line with current recommendations to detect confounding factors such as drug interactions, as well as treatment non-adherence (Ellison and Dufresne, 2015). Finally, any individual prescribed clozapine for the treatment of psychosis in Parkinson’s disease was also excluded (n = 1), leaving a final sample of 208 individuals.

### Statistical analysis of metabolite data

As a direct replication of Vaquero-Baez et al. (2019), Pearson’s correlations were computed to assess the bivariate relationships between plasma clozapine concentration, plasma norclozapine concentration, time on clozapine treatment, daily clozapine dose and ANC. Expanding on those analyses, multivariate linear regression models were used to assess the association between plasma clozapine concentration (mg/L) and ANC (cells/mm^3^) accounting for eight additional covariates: norclozapine plasma concentration (mg/L), daily clozapine dose (mg), time on clozapine treatment (days), time between clozapine concentration measurement and ANC measurement (days), time between clozapine dose and blood sampling (hours), age (years), age^2^ and sex. Following on from this baseline model, the metabolic ratio and four pharmacogenomic variants (further details below) were added to refine our assessment of the relationship between clozapine and ANC and to identify potential mediators. All statistical analyses were conducted using R v4.0.2.

### Pharmacogenomic Analyses

Four SNPs were selected for inclusion in these models, based on work conducted by Pardinas et al. (2019), in which GWAS of clozapine levels, norclozapine levels, and clozapine/norclozapine metabolic ratio were conducted to identify pharmacogenomic variants affecting the metabolism of clozapine. Details of these SNPs can be seen in **Table 1**. Another SNP reported to be associated with clozapine concentration (rs28379954; Smith et al., 2020) was also considered for analysis, but was not included due to poor imputation quality in CLOZUK2, which led to missing data in 78/208 individuals in our sample.

**Table 1:** Pharmacogenomic SNPs used in the regression analyses, and their association to clozapine metabolite concentrations in the GWAS from Pardiñas et al. 2019. * = MAF based on the European Subset of CLOZUK2 only (see Pardinas et al. 2019 for details)

| Phenotype / SNP | Minor Allele | Beta | SE | MAF* |
| --- | --- | --- | --- | --- |
| **Clozapine** |  |  |  |  |
| rs2472297 | T | -0.089 | 0.013 | 27.94 |
| **Norclozapine** |  |  |  |  |
| rs61750900 | T | -0.149 | 0.018 | 9.9 |
| rs2011425 | G | -0.112 | 0.019 | 8.65 |
| **Metabolic ratio** |  |  |  |  |
| rs61750900 | T | 0.212 | 0.012 | 9.9 |
| rs1126545 | T | 0.078 | 0.01 | 14.22 |

## Results

Descriptive statistics for the final sample can be seen in **Table 2**. This cohort represents long-term clozapine users, who at the time of sampling had ANC levels not indicative of neutropenia (ANC ≥1500 cells/mm^3^) by clozapine monitoring guidelines (Nielsen et al., 2016).

**Table 2:** Covariates used in the correlation and regression analyses, and their distribution in the CLOZUK2 sample described in this study

| **Variable** | **Average +/- SD** |  |
| --- | --- | --- |
|  | **Male N = 147** | **Female N = 61** |
| **Clozapine (mg/Litre)** | 0.47 (0.28) | 0.51 (0.32) |
| **Norclozapine (mg/Litre)** | 0.27 (0.17) | 0.29 (0.17) |
| **Daily Dose (mg/Day)** | 357 (136) | 322.2 (134) |
| **Age (Years)** | 40.3 (13.2) | 42.7 (14.2) |
| **Time on Treatment (Years)** | 3.26 (0.8) | 3.21 (1.1) |
| **Absolute Neutrophil Count (1000 Cells/mm<sup>3</sup>)** | 3.1 (1.2) | 3.1 (1.2) |

### Correlation analysis

Mirroring the approach of Vaquero-Baez et al. (2019), Pearson’s correlation statistics are reported in **Table 3**. Clozapine concentration was significantly associated with ANC in this analysis (r=-0.16; p = 0.019), whilst norclozapine, time on clozapine and daily dose were not.

**Table 3:**
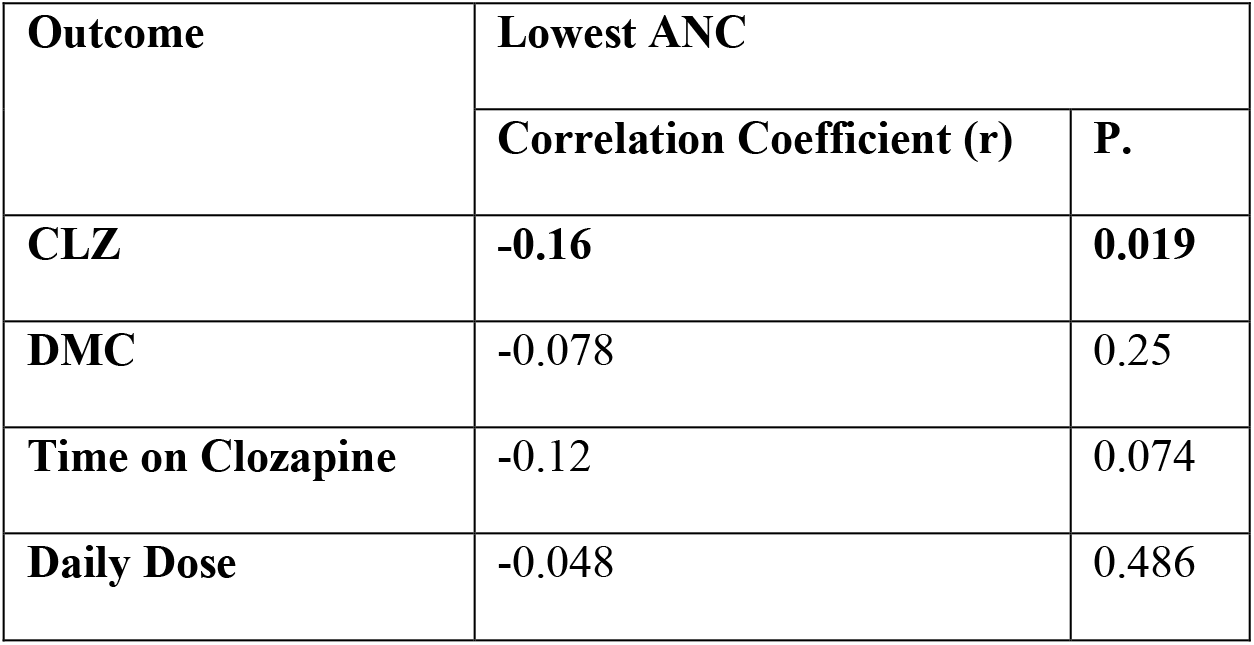
Pearson product-moment correlations of several variables with ANC, reproducing the approach of Vaquero-Baez et al. (2019)

| **Outcome** | **Lowest ANC** |  |
| --- | --- | --- |
|  | **Correlation Coefficient (r)** | **P.** |
| **CLZ** | **-0.16** | **0.019** |
| **DMC** | -0.078 | 0.25 |
| **Time on Clozapine** | -0.12 | 0.074 |
| **Daily Dose** | -0.048 | 0.486 |

### Forward stepwise regression procedure

Three regression models were considered **(Table 4)**: *Model 1* includes clozapine plasma concentration and the eight covariates described above. *Model 2* introduces the metabolic ratio as an additional covariate, following previous clozapine therapeutic drug monitoring studies (Rostami-Hodjegan et al., 2004; Couchman et al., 2013). *Model 3* additionally incorporates four genetic variants associated with clozapine metabolism (**Table 1)**

**Table 4:** Results of the three regression analyses with ANC as outcome

| **Variable** | **Model 1** |  | **Model 2** |  | **Model 3** |  |
| --- | --- | --- | --- | --- | --- | --- |
|  | *Estimates* | *p* | *Estimates* | *p* | *Estimates* | *p* |
| Clozapine Concentration | **-1.410** | **0.009** | 0.540 | 0.585 | 0.060 | 0.950 |
| Norclozapine Concentration | **1.770** | **0.049** | -1.450 | 0.385 | -0.570 | 0.738 |
| Daily Dose | 0.000 | 0.887 | 0.000 | 0.931 | 0.000 | 0.948 |
| Gender [Male] | 0.030 | 0.886 | 0.040 | 0.829 | 0.080 | 0.626 |
| Day Difference | -0.010 | 0.113 | -0.010 | 0.124 | -0.010 | 0.105 |
| Age | **0.120** | **0.001** | **0.110** | **0.001** | **0.120** | **0.001** |
| Age Squared | **-0.001** | **0.004** | **-0.001** | **0.004** | **-0.001** | **0.004** |
| Time on Clozapine | **-0.001** | **0.036** | **-0.150** | **0.044** | **-0.001** | **0.020** |
| Time Between Dose and Sample | -0.050 | 0.105 | 0.060 | 0.080 | -0.050 | 0.122 |
| Ratio |  |  | **-0.690** | **0.021** | **-0.540** | **0.035** |
| rs2472297_T (Clozapine) |  |  |  |  | -0.130 | 0.324 |
| rs61750900_T (Norclozapine + Ratio) |  |  |  |  | **-0.410** | **0.048** |
| rs2011425_G (Norclozapine) |  |  |  |  | **0.450** | **0.026** |
| rs1126545_T (Ratio) |  |  |  |  | **0.330** | **0.039** |

### Model 1

Both clozapine and norclozapine level were significantly associated with ANC; clozapine level was negatively associated (β=-1.41, p=0.009) and a positive association was found for norclozapine (β =1.77, p=0.049). For each 0.1mg/litre increase in plasma clozapine concentration, there was an associated decrease in ANC of 141 cells/mm^3^, and for each 0.1mg/litre increase in norclozapine concentration, there was an increase in ANC of 177 cells/mm^3^.

### Model 2

With the addition of metabolic ratio in the model, the effect sizes of clozapine and norclozapine shifted towards zero, becoming nonsignificant likely because of collinearity with this covariate. Thus, their initial association appeared to be explained by their ratio, which was negatively associated with ANC (β=-0.69, p=0.021). For every unit increase in the clozapine/norclozapine ratio we estimated an associated decrease of 690 cells/mm^3^ in ANC.

### Model 3

Of the four SNPs included in the analysis, 3 were significantly associated with ANC; rs61750900_T (β=-0.41, p=0.048), rs2011425_G (β=0.45, p=0.026) and rs1126545_T (β=0.33, p=0.039). Each minor allele was associated with a decrease in ANC of 410 cells/mm^3^ and increases of 450 cells/mm^3^ and 330 cells/mm^3^, respectively. In this model, the metabolic ratio remained significantly associated with a decrease in ANC (β=-0.54, p=0.035).

## Discussion

### Clozapine and neutrophil counts

Our results demonstrate a relationship between clozapine concentration and ANC, which appeared to be mediated through the metabolic ratio of clozapine and its main metabolite norclozapine. As the ratio of clozapine/norclozapine increased, ANC decreased, and this relationship appeared to be largely independent of clozapine dose. High metabolic ratios in the normal range (i.e. not suggestive of non-adherence; Ellison and Dufresne, 2015) could either be indicative of slow metabolism of clozapine or relatively rapid clearance of norclozapine, and have been previously associated to better cardiometabolic and cognitive outcomes (Costa-Dookhan et al., 2020). Regression results also highlighted a relationship with ANC of pharmacogenomic variants associated with the clozapine/norclozapine metabolic ratio, as well as norclozapine concentration. Interestingly, two of these variants (rs61750900 and rs2011425) are missense polymorphisms in UGT genes, which glucuronidise norclozapine and thereby facilitate its excretion, supporting the role of this metabolite in altering ANC (Erickson-Ridout et al., 2012). The exact biological effect that these variants might have on neutrophil count cannot be inferred from our results, though our analysis highlights a number of potential avenues for further research.

### Replication of previous results

Although our findings replicate those of Vaquero-Baez et al. (2019), the effect sizes we observed were substantially smaller. Besides the effects of winner’s curse (Kraft, 2008), and the use of a larger cohort (208 versus 41), there are a number of reasons why this may be the case. Firstly, the original study was conducted in Mexico, and the individuals recruited are likely of different ancestry (Ruiz-Linares et al., 2014) to our cohort that was recruited in the UK and made up only of those of European ancestry as inferred from genetic analyses. It cannot be ruled out that the work of Vaquero-Baez et al. (2019) has uncovered a population- or ancestry-specific effect, although no literature exists at this time to support this. An increased prevalence of neutropenia has been found in individuals with schizophrenia of African ethnicity (Kelly et al., 2007), and “benign” (constitutional) neutropenia rates vary widely based on genetic ancestry (Haddy et al., 1999; Legge et al., 2019), but no specific risk has been found in Mexican, Latino or Native American people to date.

Another noteworthy difference is that the daily clozapine dose prescribed is substantially different between the two cohorts, as inferred from a comparison of the study descriptive statistics. In our study average clozapine doses were 348 mg/day for males and 313 mg/day for females, while Vaquero-Baez et al. (2019) reported average doses of 223 mg/day for males and 105 mg/day for females (t-test p_male_=1.31 × 10^−10^; p_female_=1.43 × 10^−6^). There was also a significant difference in the average time that the cohorts had been on treatment with clozapine, upwards of three years in our study with 3.26 years for males and 3.21 years for females, and fewer than one year (Vaquero-Baez et al., 2019) with 10 months for males and 6.5 months for females (t-test p_male_=5.9 × 10^−3^; p_female_=6.18 × 10^−16^). This might have contributed to the smaller effect size we observed between clozapine metabolites and ANC, as our sample represents individuals who are long-term clozapine users and therefore likely excludes the approximately 10% of people who, after a year of treatment, might go on to exhibit neutropenia or other immune-related adverse effects (Myles et al., 2018). However, the main associations we found across our successive tests suggest that clozapine might have sustained effects on ANC even in individuals without obvious haematological adverse effects. This is consistent with the rationale for the current practice of continued haematological monitoring of people being prescribed clozapine, and supports that additional measures to lower their risk of infections might indeed be warranted even in long term clozapine users (Siskind et al., 2020), for example ensuring their timely access to the annual influenza vaccine (Pandarakalam, 2020).

### Limitations

There are a few limitations in this work that have to be noted. Although several covariates were used in the regression analyses, no data were available for the CLOZUK2 cohort for several factors that are known to affect clozapine metabolism, including concomitant medications (Singh et al., 2015), the use of tobacco (Smith and Mican, 2014) and the consumption of coffee (Raaska et al., 2004). Additionally, the cross-sectional nature of the ANC data available means there is no information regarding neutrophil trajectories throughout the individuals’ time on treatment. Replicating this analysis in a longitudinal dataset would allow for better estimating the magnitude of the detected associations and whether they vary along particular time or titration windows. This would additionally enable clarifying, through more sophisticated causal modelling, the exact relationship between clozapine metabolic ratio and ANC.

## Conclusions

Increased blood clozapine concentration is associated with decreased ANC in people with treatment-resistant schizophrenia who are long-term recipients of clozapine, even if they have not developed agranulocytosis or neutropenia during their treatment. Further investigation suggests this relationship is mediated by the clozapine/norclozapine metabolic ratio, with higher values (increased levels of clozapine proportional to norclozapine) associated with lower ANC. Furthermore, these effects can be partially explained by common genetic variants, some of which have a functional impact on enzymes involved in norclozapine glucuronidation (rs61750900 and rs2011425). Investigating these effects in a longitudinal cohort could shed further light on the relationship between clozapine and neutrophils, potentially offering biological insights. A fuller mechanistic understanding of the linkage between clozapine and ANC might allow improving the wellbeing of individuals taking clozapine by supporting targeted interventions such as prioritising seasonal vaccinations. Additional research might also lead to the elucidation of appropriate trials to assess the incorporation of genetic variants into clozapine monitoring and/or titration protocols, thus tapping into currently unrecognised but potentially valuable sources of information to improve clozapine safety at all stages of treatment.

## Data Availability

To comply with the ethical and regulatory framework of the CLOZUK project, access to individual-level data requires a collaboration agreement with Cardiff University. Requests to access data from this project should be directed to Prof. James T. R. Walters.
Genome-wide summary statistics from the GWAS cited in this study can be found at the Data Availability Link.

https://walters.psycm.cf.ac.uk/

## Acknowledgements

JTRW is supported by a UK Medical Research Council Mental Health Data Pathfinder grant (MC-PC-17212). AFP is supported by an Academy of Medical Sciences “Springboard” award (SBF005\1083). The CLOZUK2 study was supported by the following grants to Cardiff University: European Union’s Seventh Framework Programme (279227), Medical Research Council Centre (MR/L010305/1), Program (G0800509), and Project (MR/L011794/1). We acknowledge Lesley Bates, Catherine Bresner and Lucinda Hopkins for laboratory sample management at Cardiff University.

## Notes

### Competing Interest Statement

Drs. Helthuis and Jansen are full-time employees of Leyden Delta. Dr. King is a full-time employee of Magna Laboratories. Drs. Owen, O'Donovan, and Walters are supported by a collaborative research grant from Takeda Pharmaceuticals (Takeda played no part in the conception, design, implementation, or interpretation of this study). The other authors report no financial relationships with commercial interests.

### Funding Statement

Academy of Medical Sciences "Springboard" award to AFP (SBF005\1083).
MRC Mental Health Data Pathfinder grant to JTRW (MC-PC-17212).

### Author Declarations

The CLOZUK study was reviewed and approved by the UK Multicentre Research Ethics Committee (ref. 10/WSE02/15).

